# Evaluation of a home-based 7-day infection control strategy for healthcare workers following high-risk exposure to SARS-CoV-2: a cohort study

**DOI:** 10.1101/2020.11.05.20224618

**Authors:** Carla Benea, Laura Rendon, Jesse Papenburg, Charles Frenette, Ahmed Imcaoudene, Emily G McDonald, Quoc Dinh Nguyen, Ewa Rajda, Estelle Tran, Motahareh Vameghestahbanati, Andrea Benedetti, Marcel A Behr, Benjamin M Smith

## Abstract

**Background:** Evidence-based infection control strategies are needed for healthcare workers (HCWs) following high-risk exposure to SARS-CoV-2. This study evaluated the negative predictive value (NPV) of a home-based 7-day infection control strategy.

**Methods:** HCWs advised by their Infection Control or Occupational Health officer to self-isolate due to a high-risk SARS-CoV-2 exposure were enrolled between May-September 2020. The strategy consisted of symptom-triggered nasopharyngeal SARS-CoV-2 RNA testing from day 0-6 post exposure, followed by standardized home-based nasopharyngeal swab and saliva testing on day 7. The NPV of this strategy was calculated for i) clinical COVID-19 diagnosis from day 8-14 post exposure, and for ii) asymptomatic SARS-CoV-2 detected by standardized nasopharyngeal swab and saliva specimens collected at days 9-10 and 14 post exposure. Interim results are reported in the context of a second wave threatening this essential workforce.

**Results:** Among 30 HCWs enrolled to date (age 31±9 years, 24 [80.0%] female), 3 were diagnosed with COVID-19 by day 14 post exposure (secondary attack rate 10.0%), with all cases detected by the 7-day infection control strategy: NPV for subsequent clinical COVID-19 or asymptomatic SARS-CoV-2 detection by day 14 was 100.0% (95%CI: 93.1-100.0%).

**Interpretation:** Among HCWs with high-risk exposure to SARS-CoV-2, a home-based 7-day infection control strategy may have a high NPV for subsequent COVID-19 and asymptomatic SARS-CoV-2 detection. While ongoing data collection and data sharing are needed to improve the precision of the estimated NPV, we report interim results to inform infection control strategies in light of a second wave threatening this essential workforce.

## INTRODUCTION

The first wave of the COVID-19 pandemic overwhelmed healthcare systems in China, Italy, Spain, and the United States, with a second wave anticipated in Fall 2020.

High-risk exposure to SARS-CoV-2, the virus causing COVID-19, is associated with an estimated secondary attack rate of 9.1-13.8%,^1^ and exposures among healthcare workers (HCWs) during the pandemic threaten this essential workforce.^2^ Furthermore, pre- or asymptomatic SARS-CoV-2 detection following high-risk exposure has been reported,^3-11^ raising concerns of healthcare-related transmission. Evidence-based infection control strategies for HCWs following exposure to SARS-CoV-2 are needed.

Following SARS-CoV-2 exposure, the majority of secondary COVID-19 cases manifest symptoms within an estimated 5-6 days of exposure, with over 90% manifesting by day 9.^12^ Among COVID-19 cases, pre-symptomatic transmission is estimated to peak 2-3 days before symptom onset.^13^ We therefore hypothesized that a standardized home-based day 7 SARS-CoV-2 testing strategy for HCWs following high-risk exposure would have a high negative predictive value (NPV) for subsequent COVID-19 diagnosis and for asymptomatic SARS-CoV-2 detection by day 14 post exposure.

We report the interim results of a cohort study evaluating this testing strategy to provide evidence that may help inform infection control strategies in the event of a second wave.

## METHODS

### Study Design

A cohort study was initiated May 18, 2020. The study was approved by the research ethics board of the McGill University Health Centre (2020-6565), and written informed consent was obtained from all participants.

### Participants and Setting

We enrolled HCWs employed at hospital and nursing residences in the greater Montreal metropolitan area (2016 census population: 4,098,927), which had the highest COVID-19 infection rate in Canada throughout the study period (1,078 per 100,000 on May 18, 2020 to 1,433 per 100,000 on August 24, 2020).^14^

HCWs were eligible if they had a high-risk exposure to SARS-CoV-2 that required self-isolation within 7 days of eligibility screening. High-risk SARS-CoV-2 exposure requiring self-isolation was defined as i) being within 2 meters of a person with COVID-19 for at least 10 minutes without face mask or eye shield, or ii) as determined by the HCW’s institutional infection control or occupational health officer. During the study period, the participants were required to self-isolate to 14 days post exposure. Participants were not eligible if they were actively using anticoagulant medication, or unable to communicate in English or French.

### Testing strategy

Enrolled participants were advised to seek clinical testing for SARS-CoV-2 if they developed symptoms of COVID-19 (fever, significant loss of appetite, major fatigue, general muscle pain unrelated to physical exertion, sudden loss of smell, new or worsening cough, shortness of breath, sore throat, runny or stuffy nose, nausea, vomiting diarrhea, or abdominal pain) from day 0 to 7 post exposure. On day 7 post exposure, a research staff visited the participant’s self-isolation residence to collect a nasopharyngeal swab and saliva specimen for subsequent SARS-CoV-2 RNA detection by RT-PCR. The testing strategy was considered negative if SARS-CoV-2 RNA was not detected by symptom-triggered clinical testing from day 0 to 7 or the standardized home-based day 7 nasopharyngeal swab and saliva specimens.

### Outcome

The primary outcome was clinical COVID-19, defined by the presence of COVID-19 symptoms (defined above) and SARS-CoV-2 RNA detection by RT-PCR from day 8 to 14 post exposure. The secondary outcome was asymptomatic SARS-CoV-2 RNA detection by RT-PCR on standardized nasopharyngeal swab or saliva specimens, both collected on days 9-10 and 14 post exposure.

### SARS-CoV-2 RNA detection

Symptom-trigged nasopharyngeal swab specimens were collected at public testing centers and RT-PCR for SARS-CoV-2 RNA was performed within 24 hours. The home-based nasopharyngeal swab and saliva specimens collected on days 7, 9-10, and 14 post exposure were placed immediately in UTM^®^ medium and transported on ice to a −80 degree Celcius freezer within 6 hours of collection. These specimens were thawed once after a median of 74 days and SARS-CoV-2 RNA RT-PCR was performed in the clinical microbiology laboratory of the McGill University Health Centre using the cobas^®^ SARS-CoV2 Test on the Cobas 6800 System (Roche Diagnostics, Canada), the same system used for clinical care. A pilot home-collected specimen subjected to the same freeze-thaw duration confirmed detection of SARS-CoV-2 RNA was possible.

### Statistical analysis

The NPV of the testing strategy was calculated as the number of participants with a negative testing strategy result who did not experience the outcome divided by the number of participants with a negative testing strategy and expressed as a percentage. The confidence interval was calculated with an alpha of 0.05 using the binomial.confint function in the R statistical software package version 4.0.2 (www.R-project.org).

### Sample size

To inform infection control strategies in the context of a pandemic that threatens an essential workforce, we sought to maximize the precision of the lower bound of the estimated NPV confidence interval. Assuming a secondary attack rate of 10%, a testing strategy NPV of 100%, and an alpha of 0.05, enrolment of 50 participants would yield a lower bound NPV of 96%, and 100 participants would yield a lower bound NPV of 98%.

### Funding

The study was funded by the McGill Interdisciplinary Initiative in Infection and Immunity (MI4) Emergency COVID-19 Research Fund and the Rossy Foundation. The funders had no role in the study design, execution, reporting or decision to publish the findings.

## RESULTS

Interim study results are presented in the context of an anticipated imminent second wave and the lack of prospective data assessing the performance of infection control strategies for essential service providers following high-risk exposure to SARS-CoV-2.

Forty-six participants have been screened to date, and 16 were ineligible: 5 participants were outside of the 7 day post exposure enrolment period, 2 did not have a high-risk exposure, and 8 expressed interest via the online screener but did not provide contact information. One participant was eligible and consented to the participating but later withdrew consent prior to home-testing on day 7.

Among the 30 HCWs enrolled, the mean ± SD age was 31±9 years, 24 (80.0%) were female. Twenty-eight (93%) of the high-risk exposures occurred at the workplace.

A total of 151 specimens (nasopharyngeal swab and saliva) were collected by the research staff as part of the testing strategy and 30 symptom-triggered clinical tests were sought by participants. Forty-nine home-based nasopharyngeal swab and saliva specimens were collected on day 7 by the research staff (two participants were symptomatic on day 7 and sought clinical nasopharyngeal swab testing, and another provided a home-based saliva specimen but declined the nasopharyngeal swab). Eighteen symptom-triggered clinical nasopharyngeal swab tests were obtained between day 0 and 7 from 14 participants. Primary and secondary outcome ascertainment included 12 symptom-triggered clinical nasopharyngeal swab specimens collected between days 8 and 14 post exposure, and 50 standardized nasopharyngeal swab and saliva specimens collected on days 9-10, and 52 on day 14 (three and two participants declined the nasopharyngeal swab on days 9-10 and 14, respectively, but provided saliva specimens, and one participant provided a home-based nasopharyngeal swab but not a saliva specimen on day 9-10).

Three participants were diagnosed with COVID-19 by day 14 post exposure resulting in a secondary attack rate 10.0%, with all cases detected by the testing strategy (all via symptom-triggered from day 0 to 7). The NPV of the testing strategy for COVID-19 diagnosis between day 8 and 14 was 100.0% (95%CI: 93.1-100.0%) and for asymptomatic SARS-CoV-2 detection was 100.0% (95%CI: 92.3-100.0%).

## DISCUSSION

A home-based 7-day infection control strategy for HCWs following high-risk exposure to SARS-CoV-2 may have high NPV for subsequent COVID-19 and asymptomatic SARS-CoV-2 detection. Acknowledging the limited precision of the NPV estimate obtained from an interim analysis, this finding may inform local infection control policies in the event that the second wave threatens this essential workforce.

Infection control strategies for HCWs exposed to SARS-CoV-2 are guided by transmission risk and incubation time, but few have been prospectively evaluated using standardized outcome ascertainment. The present study begins to address this knowledge gap by showing that a relatively simple testing strategy may have sufficiently high NPV to allow HCWs with high-risk exposure to SARS-CoV-2 to return to work with low risk of exposure-related COVID-19 or asymptomatic SARS-CoV-2 transmission.

The present study must be considered in light of its weaknesses. First, the interim analysis resulted in a confidence interval surrounding the NPV estimate and may include unacceptably low values to merit implementation as an infection control strategy. We nevertheless present these interim results due to the lack of prospective data evaluating post exposure HCW testing strategies, and a second wave that threatens this essential workforce. With ongoing data collection and data sharing, we expect the precision of the NPV estimate to increase. Second, while the secondary attack rate in this sample is consistent with earlier close contact estimates,^1^ evolving HCW personal protective equipment policies may lower secondary attack rates in the second wave. If true, this would make our NPV estimate conservative. Third, SARS-CoV-2 immunity was not assessed but may influence the NPV of the testing strategy and the generalizability of our findings. While not yet peer-reviewed, the prevalence of SARS-CoV-2 IgG antibody detection among HCWs at a Canadian tertiary healthcare center during the same study period was low (1.4-3.4%).^15^ If immunity can be achieved through infection or vaccination, immunity will likely increase with time, making our NPV estimate conservative. Fourth, we did not perform viral culture to confirm the infectivity of participants with SARS-CoV-2 RNA detected by RT-PCR. While our pragmatic study design aimed to evaluate the testing strategy under ‘real world’ conditions (i.e., with widely available clinical tools), the assumption that any participant with SARS-CoV-2 RNA detected is infectious would make our NPV estimate conservative in terms of transmission risk. Finally, household exposure risk may vary over time, as may adherence to infection control recommendations from public health authorities. The NPV was estimated from a sample of HCWs studied during a period when schools, and businesses were closed, possibly lowering the risk of household exposure while self-isolated. Further evaluation or application of this infection control strategy should consider adherence to self-isolation and risk of secondary high-risk SARS-CoV-2 exposures.

## Conclusion

Among HCWs with high-risk exposure to SARS-CoV-2, a home-based 7-day infection control strategy may have a high NPV for subsequent COVID-19 and asymptomatic SARS-CoV-2 detection. Ongoing data collection and data sharing will maximize the precision of the estimated NPV and inform infection control strategies to protect this essential workforce.

## Data sharing

The data from this study can be made available, provided appropriate ethics approvals and data use agreements, by contacting the corresponding author. Researchers with datasets that include standardized SARS-CoV-2 RNA testing 7 days post exposure are encouraged to contact the corresponding author to conduct a individual participant data meta-analysis.

## Data Availability

The data that support the findings of this study are available from the corresponding author, Laura Rendon, upon request with appropriate ethics approvals.

